# Long-COVID post-viral chronic fatigue syndrome and affective symptoms are associated with oxidative damage, lowered antioxidant defenses and inflammation: a proof of concept and mechanism study

**DOI:** 10.1101/2022.04.25.22274251

**Authors:** Hussein Kadhem Al-Hakeim, Haneen Tahseen Al-Rubaye, Dhurgham Shihab Al-Hadrawi, Abbas F. Almulla, Michael Maes

## Abstract

The immune-inflammatory response during the acute phase of COVID-19, as assessed using peak body temperature (PBT) and peripheral oxygen saturation (SpO2), predicts the severity of chronic fatigue, depression and anxiety (“physio-affective”) symptoms three to four months later. The present study was performed to characterize whether the effects of SpO2 and PBT on the physio-affective phenome of Long COVID are mediated by immune, oxidative and nitrosative stress (IO&NS) pathways. This study assayed SpO2 and PBT during acute COVID-19, and C-reactive protein (CRP), malondialdehyde (MDA), protein carbonyls (PCs), myeloperoxidase (MPO), nitric oxide (NO), zinc, and glutathione peroxidase (Gpx) in 120 Long COVID individuals and 36 controls. Cluster analysis showed that 31.7% of the Long COVID patients had severe abnormalities in SpO2, body temperature, increased oxidative toxicity (OSTOX) and lowered antioxidant defenses (ANTIOX), and increased total Hamilton Depression (HAMD) and Anxiety (HAMA) and Fibromylagia-Fatigue (FF) scores. Around 60% of the variance in the physio-affective phenome of Long COVID (a factor extracted from HAMD, HAMA and FF scores) was explained by OSTOX/ANTIOX ratio, PBT and SpO2. Increased PBT predicted increased CRP and lowered ANTIOX and zinc levels, while lowered SpO2 predicted lowered Gpx and increased NO production. Both PBT and SpO2 strongly predict OSTOX/ATIOX during Long COVID. In conclusion, the impact of acute COVID-19 on the physio-affective symptoms of Long COVID is partly mediated by OSTOX/ANTIOX, especially lowered Gpx and zinc, increased MPO and NO production and lipid peroxidation-associated aldehyde formation. Post-viral physio-affective symptoms have an inflammatory origin and are partly mediated by neuro-oxidative toxicity.

## Introduction

Patients who have recovered from SARS-CoV-2 infection and have been discharged from hospital or outpatient settings may continue to display symptoms considerably longer than predicted ^1–5^. “Long COVID” is a blanket term that refers to people who have recovered from acute COVID-19 but are still experiencing symptoms for far longer periods than predicted ^6^. Numerous other terms for post-COVID symptoms have been suggested, including post-acute COVID, protracted COVID, chronic post-COVID, and Long Haul COVID ^7, 8^. The most often used definition is: symptoms persisting for more than three months from the onset of acute COVID-19 ^9, 10^. Extended symptoms have been classified as probably infection-related (up to 4–5 weeks), acute post-COVID symptoms (weeks 5–12), prolonged post-COVID symptoms (weeks 12–24), and chronic post-COVID symptoms (more than 24 weeks)^7^.

Following the acute phase of COVID-19, the presence of more than one symptom is quite prevalent, occurring in 74% ^11, 12^ to 87.4 percent of all infected patients ^12^. Initially, patient concerns were dismissed as mental health problems, including worry or stress ^13^, but later it became clear that people who suffer from long-term COVID have a wide range of physical and mental symptoms ^14–16^. Among the various symptoms associated with Long COVID, the most frequently reported symptoms are fatigue and dyspnoea ^9, 11, 17, 18^, post-traumatic stress symptoms ^19, 20^, concentration and memory problems ^21, 22^, and anxiety and depression ^4, 23^. Within six months after the first COVID-19 symptom, almost a third of COVID-19 survivors had a neuropsychiatric diagnosis such as insomnia, anxiety and depression ^24^.

Recently, we reported that not only the acute infectious phase ^25^ but also Long COVID ^26^ is characterized by concurrent elevations in key depression (depressed mood, feelings of guilt, suicidal ideation, loss of interest), key anxiety (anxious mood, tension, fears, anxiety behavior at interview), chronic fatigue and physiosomatic symptoms including autonomic and gastrointestinal (GIS) symptoms, malaise and muscle pain. Additionally, in both the acute phase and Long COVID, a single latent vector could be derived from these physiosomatic and affective symptoms, demonstrating that these symptom profiles are the expression of a shared core, namely the COVID-19 and Long COVID “physio-affective phenome” ^25, 26^.

We reported that in acute COVID-19, the physio-affective phenome was largely predicted by a latent factor derived from indicants of increased proinflammatory and immunoregulatory cytokines, chest computerized tomography scan abnormalities (CCTAs), including ground-glass opacities, crazy patterns and consolidation and lower oxygen saturation in peripheral blood (SpO2) ^25^. Importantly, lowered SpO2 and increased peak body temperature during the acute phase of illness largely predict the severity of the physio-affective phenome of Long COVID ^26^. Both lowered SpO2 ^26^ and increased peak body temperature ^27^ are indicants of the severity of the immune-inflammatory response of acute COVID-19, and both predict critical disease and mortality ^27, 28^. Therefore, our findings indicate that the infectious-immune-inflammatory pathways during the acute phase of illness ^29^ largely predict the physio-affective core of Long COVID ^26^. Nevertheless, there are no data on whether the biomarkers underpinning Long COVID are caused by the inflammatory responses during the acute phase.

Activated immune-inflammatory and oxidative and nitrosative stress (IO&NS) pathways may underpin the physio-affective symptoms of Long COVID because chronic fatigue syndrome, major depression and generalized anxiety disorder (GAD) are characterized by activated IO&NS pathways. Thus, these disorders are accompanied by a) signs of a mild inflammatory response as indicated by increased levels of C-reactive protein (CRP); b) oxidative stress including increased activity of pro-oxidative enzymes including myeloperoxidase (MPO) and oxidative damage as indicated by elevated production of malondialdehyde (MDA), protein carbonyls and advanced protein oxidation products (AOPP); c) increased nitrosative stress as indicated by increased levels of nitric oxide (NO) metabolites and IgM directed to NO (nitroso) neoepitope adducts; and d) indicants of lowered total antioxidant capacity (TAC) of plasma, glutathione peroxidase (Gpx) and zinc levels ^30–34^. In chronic fatigue syndrome, depression and GAD, the neurotoxicity theories conceptualize that the effects of neuro-oxidative stress toxicity (OSTOX) coupled with lowered antioxidant (ANTIOX) defenses cause increased neurotoxicity leading to physio-affective symptoms ^30, 31^.

Likewise, SARS-Cov-2 infection and acute COVID-19 are accompanied by an inflammatory response ^29^, increased MPO activity ^35^, indicants of oxidative damage ^36–38^ and increased NO production including increased levels of nitrotyrosine ^39^, and lowered antioxidant defenses as indicated by reduced TAC levels^35, 39^, Gpx ^40^ and zinc ^41^. Nevertheless, there are no data on whether the effects of the immune-inflammatory processes during acute infection, as indicated by lowered SpO2 and increased body temperature on the physio-affective phenome, are mediated by activated immune and O&NS (IO&NS) pathways.

Hence, the present study was performed to delineate whether the effects of SpO2 and peak body temperature on the physio-affective phenome of Long COVID are mediated by IO&NS pathways, including CRP, the OSTOX/ANTIOX ratio and its indicators (MDA, AOPP, carbonyl proteins, NOx, nitrotyrosine, TAC, Gpx or zinc). In addition, the present work employs the precision nomothetic approach ^31^ to define a Long COVID model which links the acute inflammation of COVID-19 with IO&NS pathways and the physio-affective phenome 3-4 months later. Moreover, we intended to construct an endophenotype class ^34^ that integrates all those biomarkers with the physio-affective phenome of Long-COVID. These findings are necessary to comprehend the biology of Long COVID and post-viral symptoms in general and may aid in predicting who will develop chronic fatigue syndrome and affective symptoms as a result of COVID-19 and viral infections in general.

## Subjects and Methods

### Subjects

We employed both a case-control research design (to explore differences between controls and Long COVID) and a retrospective cohort study design (to examine the effects of acute-phase biomarkers on Long COVID symptoms) in the current investigation. We recruited 120 patients in the last three months of 2021 who had at least two symptoms consistent with Long COVID and had previously been diagnosed and treated for acute COVID-19 infection. The patients were diagnosed using the WHO criteria of post-COVID (long COVID) ^42^, namely: a) individuals having a history of proven SARS-CoV-2 infection, b) symptoms persisted beyond the acute phase of illness or manifested during recovery from acute COVID-19 infection, c) symptoms lasted at least two months and are present 3-4 months after the onset of COVID-19, and d) patients suffer from at least two symptoms that impair daily functioning including fatigue, memory or concentration problems, shortness of breath, chest pain, persistent cough, difficulty speaking, muscle aches, loss of smell or taste, affective symptoms, cognitive impairment, or fever ^42^.

During the acute phase of the illness, all patients had been admitted to one of the official quarantine facilities in Al-Najaf city specialized in the treatment of acute COVID-19, including Al-Sader Medical City of Najaf, Al-Hakeem General Hospital, Al-Zahraa Teaching Hospital for Maternity and Pediatrics, Imam Sajjad Hospital, Hassan Halos Al-Hatmy Hospital for Transmitted Diseases, Middle Euphrates Center for Cancer, and Al-Na. Senior doctors and virologists made the diagnosis of SARS-CoV-2 infection and acute COVID 19 based on a) presence of an acute respiratory syndrome and the disease’s standard symptoms of fever, breathing difficulties, coughing, and loss of smell and taste; b) positive reverse transcription real-time polymerase chain reaction findings (rRT-PCR); and c) positive IgM directed to SARS-CoV-2. All patients showed, upon recovery from the acute phase, a negative rRT-PCR result. We selected 36 controls from the same catchment area, who were either staff members or their family or friends. Additionally, we included controls who tested negative for rRT-PCR and exhibited no clinical indications of acute infection, such as dry cough, sore throat, shortness of breath, lack of appetite, flu-like symptoms, fever, night sweats, or chills. Nevertheless, we selected the controls so that about one-third of them had distress or adjustment symptoms as a result of lockdowns and social isolation to account for their confounding effects, which are also seen in Long COVID patients. Thus, one-third of the controls showed Hamilton Depression Rating Scale (HAMD) ^43^ values between 7 and 12. COVID patients and controls were excluded if they had a lifetime history of psychiatric axis-1 disorders, including major affective disorders such as major depression and bipolar disorder, dysthymia, generalized anxiety disorder and panic disorder, schizo-affective disorder, schizophrenia, psycho-organic syndrome, and substance use disorders, except tobacco use disorder (TUD). Moreover, we excluded patients and controls who suffered from neurodegenerative and neuroinflammatory disorders, such as chronic fatigue syndrome ^44^, Parkinson’s or Alzheimer’s disease, multiple sclerosis, stroke, or systemic (auto)immune diseases such as diabetes mellitus, psoriasis, rheumatoid arthritis, inflammatory bowel disease and scleroderma, and liver and renal disease. Additionally, pregnant and breastfeeding women were omitted from this study.

All controls and patients, or their parents/legal guardians, gave written informed permission before participation in the research. The research was approved by the University of Kufa’s institutional ethics board (8241/2021) and the Najaf Health Directorate-Training and Human Development Center (Document No.18378/ 2021). The study was conducted following Iraqi and international ethical and privacy laws, including the World Medical Association’s Declaration of Helsinki, The Belmont Report, the CIOMS Guideline, and the International Conference on Harmonization of Good Clinical Practice; our institutional review board adheres to the International Guidelines for Human Research Safety (ICH-GCP).

#### Clinical measurements

A senior psychiatrist used a semi-structured interview to collect socio-demographic and clinical data from controls and Long COVID patients three to four months after the acute infectious phase of COVID-19 (mean ±SD duration of illness was 14.68 ±5.31 weeks). Three to four months after the onset of acute COVID-19, a senior psychiatrist assessed the following rating scales: a) the 12-item Fibro-Fatigue (FF) scale to assess Chronic fatigue and fibromyalgia symptoms ^45^; b) the HAMD to assess the severity of depression ^43^; and c) the Hamilton Anxiety Rating Scale (HAMA) ^46^ to assess the severity of anxiety. Two HAMD subdomain scores were calculated: a) pure depressive symptoms (pure HAMD) as the sum of sad mood + feelings of guilt + suicidal thoughts + loss of interest; and b) physiosomatic HAMD symptoms (Physiosom HAMD) as the sum of somatic anxiety + gastrointestinal (GIS) anxiety + genitourinary anxiety + hypochondriasis. Two HAMA subdomain scores were calculated: a) key anxiety symptoms (Key HAMA), which were defined as anxious mood + tension + fears + anxiety behavior during the interview; and b) physiosomatic HAMA symptoms (Physiosom HAMA), defined as somatic sensory + cardiovascular + GIS + genitourinary + autonomic symptoms (respiratory symptoms were not included in the sum). We calculated a single pure physiosom FF subdomain score as: muscular pain + muscle tension + fatigue + autonomous symptoms + gastrointestinal symptoms + headache + a flu-like malaise (thus excluding the cognitive and affective symptoms). To construct a composite score reflecting the severity of the physio-affective phenome, we extracted the first factor from the pure FF and pure and physiosom HAMA and HAMD scores, which reflect the physio-affective phenome ^25, 26^. Additionally, we constructed z unit-based composite scores indicating autonomic symptoms, sleep problems, fatigue, gastro-intestinal symptoms, and cognitive symptoms using all relevant HAMD, HAMA, and FF items (z transformed). TUD was diagnosed using DSM-5 criteria. We determined the body mass index (BMI) by dividing the body weight in kilograms by the squared height in meters.

#### Assays

Fasting blood samples were taken in the early morning between 7.30-9.00 a.m. after awakening and before having breakfast. Five milliliters of venous blood samples were drawn and transferred into clean plain tubes. Hemolyzed samples were rejected. After ten minutes, the clotted blood samples were centrifuged for five minutes at 3000 rpm, and then serum was separated and transported into three new Eppendorf tubes until assay. Serum Gpx, NOx, MPO, MDA, AOPP, TAC, nitrotyrosine, and protein carbonyls were measured using ELISA kits supplied by Nanjing Pars Biochem Co., Ltd. (Nanjing, China). All kits were based on a sandwich technique and showed an inter-assay CV of less than 10%. Zinc in serum was measured spectrophotometrically using a ready for use kit supplied by Agappe Diagnostics Ltd., Cham, Switzerland. Consequently, we computed 3 composite scores: a) oxidative stress toxicity (OSTOX) as the sum of z MDA + z AOPP + z carbonyl proteins + z MPO + z NOx + z nitrotyrosine; b) antioxidant defenses (ANTIOX) as z TAC + z Gpx + z zinc; and c) the OSTOX/ANTIOX ratio as z OSTOX – z ANTIOX.

A well-trained paramedical specialist measured SpO2 using an electronic oximeter supplied by Shenzhen Jumper Medical Equipment Co. Ltd., and a digital thermometer was used to assess body temperature (sublingual until the beep). We collected these indicators from patient records and analyzed the lowest SpO2 and peak body temperature values recorded during the acute phase of illness. We created a new indicator based on these two evaluations that represent decreased SpO2 and increased peak body temperature as the z transformation of the latter (z body temperature) - z SpO2 (dubbed the “TO2 index”). We recorded the immunizations received by each participant, namely AstraZeneca, Pfizer, or Sinopharm.

#### Statistics

Analysis of variance (ANOVA) was performed to determine if there were variations in scale variables across groups, and analysis of contingency tables was employed to determine connections between nominal variables. We used Pearson’s product-moment correlation coefficients to examine relationships between ONS biomarkers and SpO2, peak body temperature, and clinical physio-affective assessments. We employed a univariate general linear model (GLM) analysis to characterize the associations between classifications and biomarkers while accounting for confounding factors such as TUD, sex, age, BMI, and education. Additionally, we obtained model-generated estimated marginal means (SE) values and used protected pairwise comparisons between group means using Fisher’s least significant difference. Multiple comparisons were examined using a p-correction for false discovery rate (FDR) ^47^. Multiple regression analysis was used to determine the important IO&NS biomarkers or cofounders that predict physio-affective measures. We used an automated stepwise technique with a 0.05 p-value to enter and p=0.06 to remove. We calculated the standardized beta coefficients for each significant explanatory variable using t statistics with exact p-value, as well as the model F statistics and total variance explained (R^2^). Additionally, we examined multicollinearity using the variance inflation factor and tolerance. We checked for heteroskedasticity using the White and modified Breusch-Pagan tests, and where necessary, we generated parameter estimates with robust errors. The tests were two-tailed, and statistical significance was defined as a p-value of 0.05. Using an effect size of 0.23, a p-value of 0.05, a power of 0.8, and three groups with up to five variables in an analysis of variance, the sample size should be around 151 participants. As a result, we enrolled 156 individuals, 36 controls and 120 Long COVID participants.

#### The precision nomothetic approach

By integrating biomarker and clinical data, we hoped to create endophenotype classes for Long COVID patients (using cluster analysis) and novel pathway phenotypes (using factor analysis). We conducted two-step cluster analyses on categorical (e.g., diagnosis) and scale variables (e.g., all biomarkers) to define new meaningful clusters of Long COVID patients. When the silhouette measure of cohesiveness and separation was more than 0.4, the cluster solution was deemed satisfactory. We conducted exploratory factor analysis (unweighted least-squares extraction, 25 iterations for convergence) and assessed factorability using the Kaiser-Meier-Olkin (KMO) sample adequacy metric (adequate when >0.7). Additionally, when all loadings on the first factor were greater than 0.6, the variance explained by the first factor was greater than 50.0 percent, and the Cronbach alpha on the variables was greater than 0.7; the first factor was considered a validated latent construct underlying the variables. Canonical correlation analysis (CCA) was used to investigate the associations between two sets of variables, with symptoms three to four months following the acute phase serving as the dependent variable set and both biomarkers of the acute and Long COVID phases as the explanatory variable set. We calculated the variance explained by the canonical variables in both sets, and we accept the canonical sets as an overall measure of the underlying construct when the explained variance is > 0.50 and all canonical loadings are > 0.5. Finally, we also compute the variance explained in the dependent clinical set by the biomarker set. All statistical analyses were conducted using IBM SPSS Windows, version 28.

Smart PLS analysis ^48^ was utilized to investigate the causal links between the lowest SpO2 and peak body temperature in the acute phase and the physio-affective phenome in Long COVID, whereby the effects of SpO2 and temperature are mediated by IO&NS biomarkers. All input variables were entered as single indicators, while the output variable was a latent vector extracted from pure FF and pure and physiosom HAMD and HAMA values. Complete PLS analysis was performed only when the inner and outer models met predefined quality criteria, namely: a) the output LV has high composite reliability >0.7, Cronbach’s alpha >0.7, and rho A >0.8 with an average variance extracted (AVE) > 0.5, b) all LV loadings are > 0.6 at p < 0.001, c) the model fit is < 0.08 in terms of standardized root mean squared residual (SRMR), d) confirmatory tetrad analysis shows that the LV was not incorrectly specified as a reflective model, e) blindfolding shows that the construct’s cross-validated redundancy is adequate, and f) the model’s prediction ability as evaluated using PLSPredict is adequate. If the model quality data are adequate, we perform a complete PLS pathway analysis using 5000 bootstrap samples and compute the path coefficients with exact p-value, as well as specific and total indirect (mediated) effects and total effects.

## Results

### Construction of an endophenotype class based on all biomarkers

To discover new endophenotype classes of Long COVID patients, we used a two-step cluster analysis with the diagnosis (Long COVID versus controls as a category) and the acute COVID-19 biomarkers SpO2 and body temperature and Long COVID biomarkers OSTOX, ANTIOX, OSTOX/ANTIOX and CRP as continuous variables. Nevertheless, the solution without CRP was much better, and, therefore, CRP was deleted from the solution. We found a three-group model with an adequate measure of cohesion and separation of 0.57, namely controls (n=36) and two patient clusters comprising 67 (cluster 1) and 51 (cluster 2) patients, respectively. Table 1 shows the socio-demographic and biomarker data of the three clusters. Cluster 2 patients showed lower SpO2, zinc, Gpx and ANTIOX levels and higher body temperature, OSTOX, OSTOX/ANTIOX, NOx, MPO, MDA and protein carbonyl levels as compared with cluster 1 patients. As such, the Long COVID group is divided into two clusters, one (cluster 2) with highly activated O&NS pathways (LC+O&NS) and one with less severe aberrations (LC). The LC group was differentiated from controls by increased body temperature and CRP and lowered SpO2, Gpx and ANTIOX values.

**Table 1:** Socio-demographic and biomarkers data of controls and Long COVID (LC) patients divided into two groups one with highly increased body temperature, lowered SpO2 and high nitro-oxidative stress (LC+O&NS) versus another group with fewer changes in these biomarkers (LC).

| Variables | Controls<br>(n=36) <sup>A</sup> | Cluster 1 or<br>LC (n=67) <sup>B</sup> | Cluster 2: LC +<br>O&NS (n=51) <sup>C</sup> | F/$\psi$/$\chi^2$ | df | p |
| --- | --- | --- | --- | --- | --- | --- |
| Age (years) | 30.9 ± 8.3 | 29.3 ± 7.2 | 33.1 ± 8.6 | 3.32 | 2/151 | 0.039 |
| BMI (kg/m<sup>2</sup>) | 26.34 ± 3.64 | 25.60 ± 3.97 | 26.65 ± 6.20 | 0.72 | 2/146 | 0.487 |
| Sex (Female/Male) | 6/30 | 25/42 | 8/43 | 9.02 | 2 | 0.012 |
| Education (years) | 15.8 ± 1.2 <sup>C</sup> | 15.6 ± 1.6 <sup>C</sup> | 15.5 ± 1.8 <sup>A,B</sup> | 0.37 | 2/151 | 0.689 |
| Single/married | 22/14 | 34/33 | 15/36 | 9.56 | 2 | 0.008 |
| TUD (No/Yes) | 20/16 | 45/22 | 36/15 | 2.24 | 2 | 0.321 |
| Residency (Urban/Rural) | 7/29 | 7/60 | 12/39 | 3.75 | 2 | 0.166 |
| Employment (No/Yes) | 36 | 67 | 51 | - | - | - |
| Vaccination (A/Pf/S) | 11/14/11 | 23/30/14 | 20/19/12 | 1.83 | 4 | 0.768 |
| Duration of Disease (weeks) | - | 13.8 ± 3.7 <sup>C</sup> | 15.8 ± 6.8 <sup>B</sup> | 4.16 | 1/116 | 0.044 |
| Peak body temperature (°C) | 36.5 ± 0.1 <sup>B,C</sup> | 38.8 ± 0.1 <sup>A,C</sup> | 39.3 ± 0.1 <sup>A,B</sup> | 163.99 | 2/142 | <0.001 |
| Lowest SpO2 (%) | 96.6 ± 0.6 <sup>B,C</sup> | 92.3 ± 0.4 <sup>A,C</sup> | 87.5 ± 0.5 <sup>A,B</sup> | 70.06 | 2/142 | <0.001 |
| zTO2 index (z scores) | -1.340 ± 0.091 <sup>B,C</sup> | 0.143 ± 0.070 <sup>A,C</sup> | 0.862 ± 0.079 <sup>A,B</sup> | 170.24 | 2/142 | <0.001 |
| Gpx (ng/ml) | 23.46 ± 0.97 <sup>B,C</sup> | 21.57 ± 0.75 <sup>A,C</sup> | 17.97 ± 0.84 <sup>A,B</sup> | 9.84 | 2/142 | <0.001 |
| NO (uM) | 20.81 ± 1.47 <sup>C</sup> | 18.95 ± 1.13 <sup>C</sup> | 28.19 ± 1.27 <sup>A,B</sup> | 14.99 | 2/142 | <0.001 |
| Protein carbonyl (ng/ml) | 11.38 ± 1.15 <sup>C</sup> | 10.73 ± 0.89 <sup>C</sup> | 14.43 ± 1.00 <sup>A,B</sup> | 3.97 | 2/142 | 0.021 |
| MPO (ng/ml) | 12.87 ± 0.82 <sup>C</sup> | 11.25 ± 0.63 <sup>C</sup> | 15.84 ± 0.71 <sup>A,B</sup> | 11.32 | 2/142 | <0.001 |
| MDA (nM) | 1406.9 ± 97.9 <sup>C</sup> | 1432.9 ± 75.5 <sup>C</sup> | 1843.0 ± 84.6 <sup>A,B</sup> | 8.02 | 2/142 | <0.001 |
| AOPP (uM) | 4.84 ± 0.38 | 5.27 ± 0.29 | 5.87 ± 0.33 | 2.19 | 2/142 | 0.116 |
| TAC (U/ml) | 6.23 ± 0.48 | 5.23 ± 0.37 | 5.01 ± 0.41 | 2.09 | 2/142 | 0.128 |
| Nitrotyrosine (nM) | 33.44 ± 3.20 | 34.79 ± 2.46 | 32.72 ± 2.76 | 0.16 | 2/142 | 0.865 |
| Zinc (ppm) | 0.785 ±0.034 <sup>C</sup> | 0.772 ±0.026 <sup>C</sup> | 0.657 ±0.030 <sup>A,B</sup> | 5.37 | 2/142 | 0.006 |
| OSTOX (z scores) | -0.357 ±0.131 <sup>C</sup> | -0.512 ±0.101 <sup>C</sup> | 0.882 ±0.113 <sup>A,B</sup> | 45.19 | 2/142 | <0.001 |
| ANTIOX (z scores) | 0.521 ±0.151 <sup>B,C</sup> | 0.107 ±0.116 <sup>A,C</sup> | -0.580 ±0.130 <sup>A,B</sup> | 16.17 | 2/142 | <0.001 |
| OSTOX / ANTIOX (z scores) | -0.575 ±0.129 <sup>A</sup> | -0.406 ±0.100 <sup>A</sup> | 0.957 ±0.113 <sup>A,B</sup> | 53.62 | 2/142 | <0.001 |
| CRP (mg/L) | 5.42 (0.70) <sup>B,C</sup> | 7.83 (0.55) <sup>A</sup> | 9.11 (0.61) <sup>A</sup> | 7.96 | 2/140 | <0.001 |

All results are shown as mean (SD) or as ratios. <sup>A,B,C</sup> post-hoc comparisons among group means
BMI: body mass index, TUD: tobacco-use disorder, Vaccination A1,Pf,S: vaccination type either AstraZeneca (A), Pfizer-BioNTech (Pf), or Sinopharm (S).
zTO2: composite score reflecting combined aberrations in body temperature and SpO2, Gpx: glutathione peroxidase, NOx: nitric oxide, MPO: myeloperoxidase, MDA: malondialdehyde, AOPP: advances oxidation protein products, TAC: total antioxidant capacity.

**Table 1** demonstrates the socio-demographic data in Long COVID patients divided into LC and LC+O&NS. No significant differences among these study groups were detected in BMI, residency, employment, education, vaccination status, and TUD. Age was somewhat higher in the LC+O&NS group, and there were more males in the LC+O&NS group than in the LC group. The disease duration was somewhat higher in LC+O&NS as compared with LC.

### Associations between LC clusters and physio-affective symptoms of Long COVID

**Table 2** shows the results of univariate GLM analysis examining the associations between diagnosis into controls, LC and LC+IO&NS and all clinical scores while controlling for the effects of age, sex, education years, and TUD (entered as covariates). We found that the total FF, HAMD and HAMA, pure FF, and pure and physiosom HAMD and HAMA scores, as well as the autonomic and GIS symptoms, were significantly different between the three classes and increased from controls → LC → LC+O&NS. Sleep disorders, chronic fatigue and cognitive disorders were significantly higher in Long Covid than in controls.

**Table 2.** Estimated marginal physio-affective mean scores in controls and Long COVID patients divided into two groups one with highly increased body temperature, lowered SpO2 and high nitro-oxidative stress (LC+O&NS) versus another group with fewer changes in these biomarkers (LC).

| **Biomarkerse** | **Controls <sup>A</sup>** | **Cluster 1 or LC (n=67)** | **Cluster 2: LC +O&NS (n=51)** | **F** | **df** | **p** |
| --- | --- | --- | --- | --- | --- | --- |
| Total HAMD | 6.01 (0.82) <sup>B,C</sup> | 15.24 (0.61) <sup>A,C</sup> | 20.17 (0.70) <sup>A,B</sup> | 88.39 | 2/147 | <0.0001 |
| Total HAMA | 7.65 (1.27) <sup>B,C</sup> | 17.78 (0.94) <sup>A,C</sup> | 23.72 (1.08) <sup>A,B</sup> | 46.95 | 2/147 | <0.0001 |
| Total FF | 7.11 (1.66) <sup>B,C</sup> | 23.30 (1.24) <sup>A,C</sup> | 31.06 (1.42) <sup>A,B</sup> | 61.66 | 2/147 | <0.0001 |
| Pure HAMD | 1.65 (0.29) <sup>B,C</sup> | 4.22 (0.21) <sup>A,C</sup> | 5.85 (0.25) <sup>A,B</sup> | 62.39 | 2/147 | <0.0001 |
| Psysiosom HAMD | 1.65 (0.34) <sup>B,C</sup> | 4.34 (0.26) <sup>A,C</sup> | 5.92 (0.29) <sup>A,B</sup> | 44.95 | 2/147 | <0.0001 |
| Pure HAMA | 1.65 (0.33) <sup>B,C</sup> | 3.38 (0.25) <sup>A</sup> | 4.06 (0.28) <sup>A</sup> | 16.26 | 2/147 | <0.0001 |
| Physiosom HAMA | 3.17 (0.61) <sup>B,C</sup> | 7.86 (0.46) <sup>A,C</sup> | 11.52 (0.53) <sup>A,B</sup> | 53.57 | 2/147 | <0.0001 |
| Pure FF | 4.61 (1.01) <sup>B,C</sup> | 15.69 (0.76) <sup>A,C</sup> | 20.71 (0.87) <sup>A,B</sup> | 74.64 | 2/147 | <0.0001 |
| Physio-affective phenome (z score) | -1.167 (0.118) <sup>B,C</sup> | 0.043 (0.088) <sup>A,C</sup> | 0.766 (0.101) <sup>A,B</sup> | 77.78 | 2/147 | <0.001 |
| Autonomic symptoms (z score) | -1.157 (0.122) <sup>B,C</sup> | 0.139 (0.091) <sup>A,C</sup> | 0.623 (0.104) <sup>A,B</sup> | 64.53 | 2/147 | <0.0001 |
| Sleep disorders (z score) | -0.936 (0.141) <sup>B,C</sup> | 0.022 (0.105) <sup>A</sup> | 0.591 (0.121) <sup>A</sup> | 34.10 | 2/147 | <0.0001 |
| Fatigue (z score) | -1.036 (0.136) <sup>B,C</sup> | 0.167 (0.102) <sup>A</sup> | 0.519 (0.117) <sup>A</sup> | 40.42 | 2/147 | <0.0001 |
| Gastro-intestinal symptoms (z score) | -0.915 (0.143) <sup>B,C</sup> | 0.117 (0.107) <sup>A,C</sup> | 0.474 (0.123) <sup>A,B</sup> | 28.59 | 2/147 | <0.0001 |
| Cognitive disorders (z score) | -0.532 (0.163) <sup>B,C</sup> | 0.074 (0.122) <sup>A</sup> | 0.257 (0.140) <sup>A</sup> | 7.22 | 2/147 | 0.002 |

All results are shown as mean (SD) or as ratios. <sup>A,B,C</sup> posthoc comparisons among group means
Physiosom: physiosomatic; FF: Fibro Fatigue scale; HAMD: Hamilton Depression Rating Scale; HAMA: Hamilton Anxiety Rating Scale;
Physio-affective phenome: z unit-based composete score based on FF, HAMD and HAMA scores

**Table 3** shows the intercorrelation matrix of the O&NS biomarkers, on the one hand, and SpO2, maximal body temperature, and clinical physio-affective ratings, on the other. OSTOX was significantly associated with SpO2, all physio-affective ratings, except cognitive impairment scores. There was no significant association between OSTOX and body temperature. ANTIOX was inversely correlated with all physio-affective ratings, including cognitive impairments and was positively associated with SpO2. The OSTOX /ANTIOX ratio was significantly associated with all physio-affective scores, SpO2 and body temperature. There were no significant associations between CRP and the OSTOX/ANTIOX measurements. **Figure 1** shows the partial regression of the physio-affective phenome score on OSTOX/ANTIOX after adjusting for age, sex, BMI, education and TUD.

**Figure 1.**
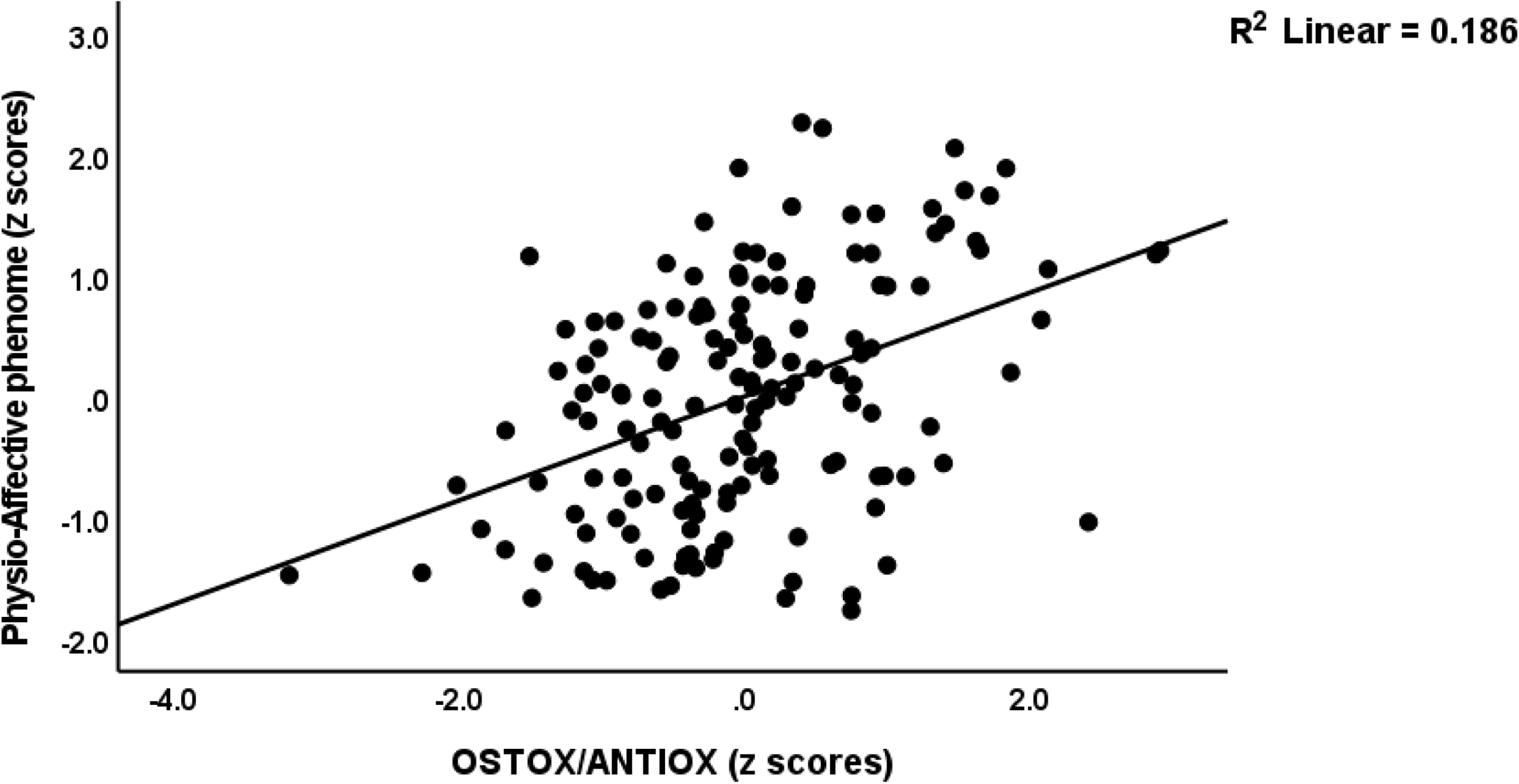
Partial regression of the physio-affective phenome score on the oxidative stress toxicity / antioxidant (OSTOX/ANTIOX) ratio in patients with Long COVID

**Table 3.** Intercorrelations between oxidative stress toxicity (OSTOX), antioxidant defenses (ANTIOX), the OSTOX/ANTIOX ratio and oxygen saturation (SpO2), peak body temperature, C-reactive protein (CRP), and physio-affective rating scale scores

| Variables | OSTOX | ANTIOX | OSTOX /ANTIOX | BTO2ONS index |  |
| --- | --- | --- | --- | --- | --- |
| SpO2 | -0.279 | 0.343 | -0.405 |  |  |
| Body temperature | 0.156 <sup>\$</sup> | -0.270 | 0.277 | | |
| CRP | 0.088 <sup>\$</sup> | -0.102 <sup>\$</sup> | 0.123 <sup>\$</sup> | 0.432 | 0.391 |
| Total HAMD | 0.308 | -0.287 | 0.388 | 0.710 | 0.516 |
| Total HAMA | 0.249 | -0.341 | 0.384 | 0.677 | 0.495 |
| Total FF | 0.322 | -0.318 | 0.417 | 0.705 | 0.502 |
| Pure HAMD | 0.344 | -0.226 | 0.371 | 0.604 | 0.342 |
| Psysiosom HAMD | 0.293 | -0.300 | 0.386 | 0.634 | 0.427 |
| Pure HAMA | 0.157 * | -0.223 | 0.248 | 0.472 | 0.270 |
| Physiosom HAMA | 0.286 | -0.336 | 0.405 | 0.711 | 0.570 |
| Pure FF | 0.334 | -0.306 | 0.417 | 0.732 | 0.535 |
| Physio-affective phenome (z score) | 0.330 | -0.338 | 0.435 | 0.752 | 0.574 |
| Autonomic symptoms (z score) | 0.283 | -0.313 | 0.388 | 0.713 | 0.515 |
| Sleep disorders (z score) | 0.247 | -0.210 | 0.298 | 0.573 | 0.391 |
| Fatigue (z score) | 0.222 | -0.364 | 0.382 | 0.622 | 0.392 |
| GIS (z score) | 0.177 * | -0.189 | 0.238 | 0.514 | 0.306 |
| Cognitive disorders (z score) | 0.136 <sup>\$</sup> | -0.224 | 0.234 | 0.338 | 0.202 <sup>\$</sup> |
| n | n=156 | n=156 | n=156 | n=156 | n=120 |

All at $p < 0.01$ , except \* $p < 0.05$ and <sup>\$</sup> nonsignificant. FF: Fibro Fatigue scale, HAMD: Hamilton Depression Rating Scale, HAMA: Hamilton Anxiety Rating Scale, GIS: gastrointestinal system, BTO2ONS: a z unit based composite score based on body temperature, SpO2, OSTOX and ANTIOX. The latter was computed in the total and the restricted sample of Long COVID patients.

### Prediction of the physio-affective scores by IO&NS biomarkers

**Table 4** shows the results of multiple regression analyses with physio-affective measurements as dependent variables and O&NS biomarkers and CRP as explanatory variables while allowing for the effects of age, sex, BMI, TUD, education and vaccination types. Regression #1 shows that 25.0% of the variance in the pure HAMD score could be explained by the regression on Gpx and education (inversely associated) and MDA, CRP and carbonyl proteins (positively associated). We found that 23.0% of the variance in the physiosom HAMD score (regression #2) was explained by the cumulative effects of lowered Gpx and zinc, increased NO and CRP and vaccination with AstraZeneca or Pfizer. Regression #3 shows that 13.1% of the variance in pure HAMA is explained by CRP, female sex and AstraZeneca vaccination. In Regression #4, 26.7% of the variance in physiosom HAMA was explained by the regression on CRP and NO (positively) and Gpx (inversely), female sex and vaccination with AstraZeneca or Pfizer. Regression #5 shows that 28.0% of the variance in pure FF was explained by NO, MDA, CRP (positively) and Gpx (inversely). Up to 30.2% of the variance in the severity of the physio-affective phenome score was explained by NO, MDA, CRP (positively), Gpx (inversely), female sex, and vaccination with AstraZeneca or Pfizer.

**Table 4.**
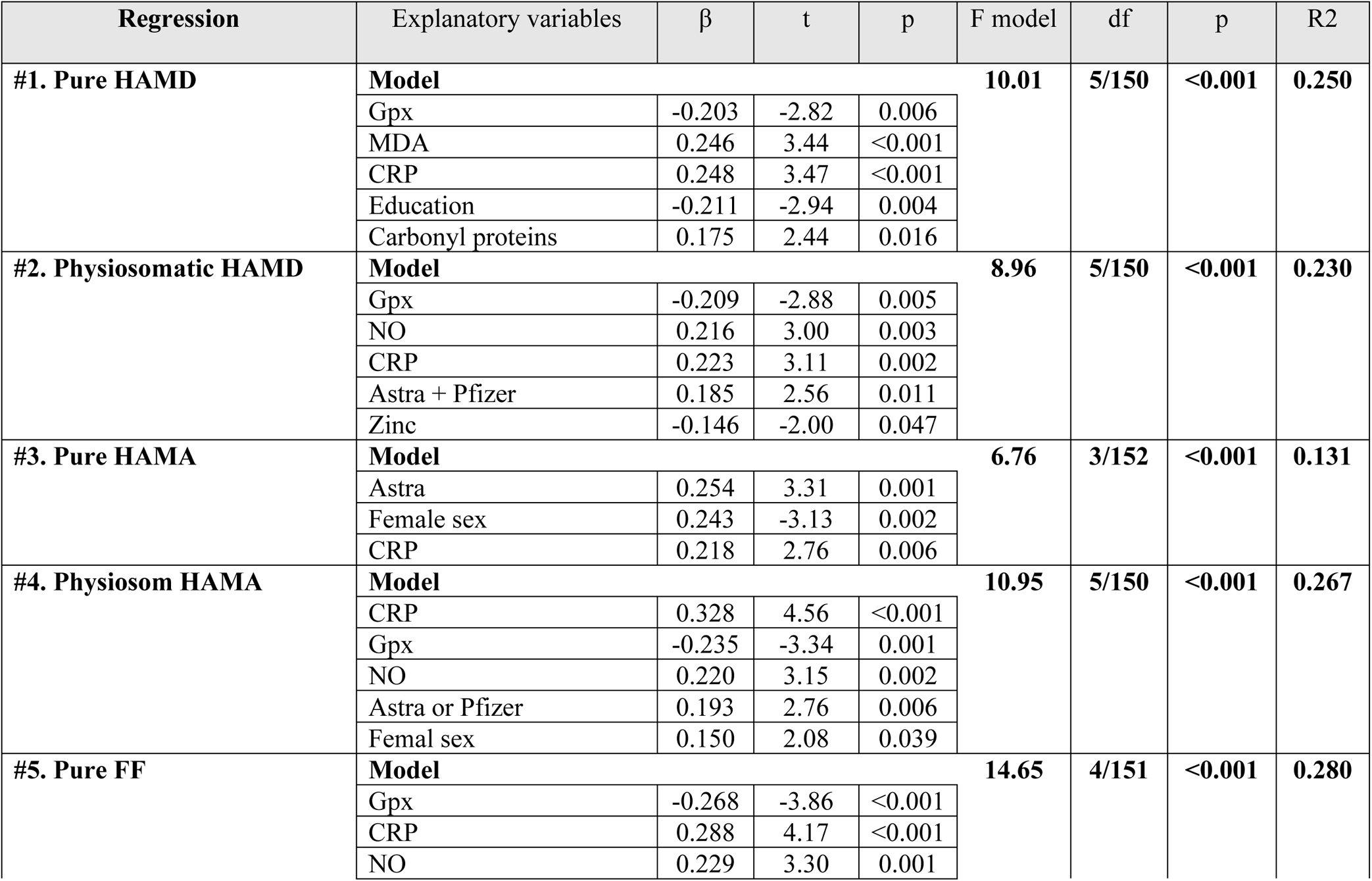
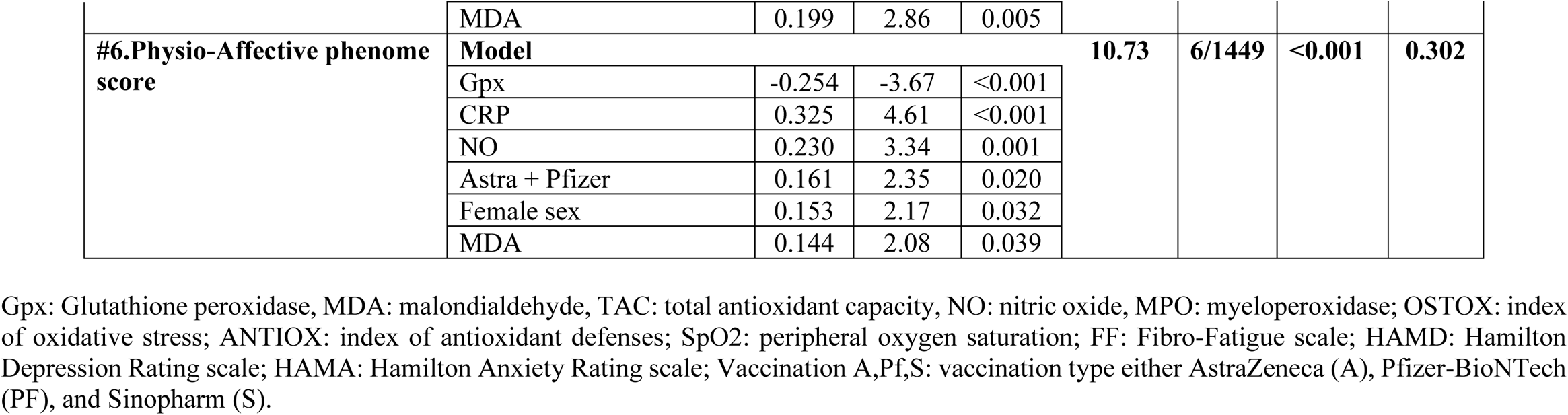
Results of multiple regression analyses with physio-affective measurements as dependent variables and immune and oxidative and nitrosative stress biomarkers as explanatory variables

### Prediction of physio-affective scores by IO&NS biomarkers, SpO2 and body temperature

We have rerun the analyses shown in Table 4 and include the OSTOX, ANTIOX, OSTOX/ANTOX ratio, SpO2 and body temperature and the results are presented in **Table 5**. SpO2 was inversely associated with all 6 physio-affective scores (regressions #1 - #6), body temperature was positively associated with all scores, except pure HAMA, while IO&NS biomarkers had significant effects on all scores above and beyond the effects of SpO2 and body temperature (all except pure HAMA). Thus, OSTOX was associated with pure HAMD, NO with physiosom HAMD and HAMA and pure FF, MDA predicted pure FF and OSTOX/ANTIOX predicted the physio-affective phenome. The effects of CRP disappeared after considering the effects of the other explanatory variables.

**Table 5.** Results of multiple regression analyses with physio-affective measurements as dependent variables and oxidative stress toxicity (OSTOX) and antioxidant (ANTIOX) biomarkers as explanatory variables

| Regression | Explanatory variables | $\beta$ | t | p | F model | df | p | R<sup>2</sup> |
| --- | --- | --- | --- | --- | --- | --- | --- | --- |
| #1. Pure HAMD | **Model** |  |  |  | **23.16** | **5/150** | **<0.001** | **0.441** |
|  | Body temperature | 0.390 | 5.12 | <0.001 |  |  |  |  |
|  | OSTOX | 2.37 | 3.70 | <0.001 |  |  |  |  |
|  | SpO2 | -0.195 | -2.49 | 0.014 |  |  |  |  |
|  | Education | -0.161 | -2.59 | 0.011 |  |  |  |  |
|  | Age | -0.137 | -2.21 | 0.028 |  |  |  |  |
| #2. Physiosom HAMD | **Model** |  |  |  | **30.59** | **4/151** | **<0.001** | **0.448** |
|  | SpO2 | -0.427 | -5.48 | <0.001 |  |  |  |  |
|  | Body temperature | 0.238 | 3.14 | 0.002 |  |  |  |  |
|  | Astra + Pfizer | 0.158 | 2.57 | 0.011 |  |  |  |  |
|  | NO | 0.149 | 2.41 | 0.017 |  |  |  |  |
| #3. Pure HAMA | **Model** |  |  |  | **26.14** | **3/152** | **<0.001** | **0.340** |
|  | SpO2 | -0.511 | -7.64 | <0.001 |  |  |  |  |
|  | Female sex | 0.223 | -3.38 | <0.001 |  |  |  |  |
|  | Astra + Pfizer | 0.133 | 1.99 | 0.048 |  |  |  |  |
| #4. Physiosom HAMA | **Model** |  |  |  | **41.14** | **5/150** | **<0.001** | **0.578** |
|  | SpO2 | -0.533 | -7.78 | <0.001 |  |  |  |  |
|  | Body temperature | 0.241 | 3.61 | <0.001 |  |  |  |  |
|  | Astra + Pfizer | -0.134 | 2.50 | 0.014 |  |  |  |  |
|  | NO | 0.123 | 2.27 | 0.025 |  |  |  |  |
|  | Femal sex | -0.114 | -2.13 | 0.035 |  |  |  |  |
| #5. Pure FF | **Model** |  |  |  | **53.60** | **4/151** | **<0.001** | **0.587** |
|  | SpO2 | -0.457 | -6.84 | <0.001 |  |  |  |  |
|  | Body temperature | 0.322 | 4.95 | <0.001 |  |  |  |  |
|  | NO | 0.151 | 2.80 | 0.006 |  |  |  |  |
|  | MDA | 0.132 | 2.49 | 0.014 |  |  |  |  |
| #6. Physio-Affective phenome score | **Model** |  |  |  | **59.77** | **4/151** | **<0.001** | **0.613** |
|  | SpO2 | -0.531 | -8.02 | <0.001 |  |  |  |  |
|  | Body temperature | 0.246 | 3.90 | <0.001 |  |  |  |  |
|  | OSTOX/ANTIOX | 0.151 | 2.73 | 0.007 |  |  |  |  |
|  | Female sex | 0.111 | 2.19 | 0.030 |  |  |  |  |
| #7. OSTOX/ANTIOX | **Model** |  |  |  | **30.26** | **1/154** | **<0.001** | **0.164** |
|  | SpO2 | -0.405 | -5.50 | <0.001 |  |  |  |  |
| #9. CRP | **Model** |  |  |  | **46.14** | **2/153** | **<0.001** | **0.376** |
|  | Body Temperature | 0.568 | 8.90 | <0.001 |  |  |  |  |
|  | Male sex | 0.245 | 3.83 | <0.001 |  |  |  |  |

Gpx: Glutathione peroxidase, MDA: malondialdehyde, NO, nitric oxide,,: SpO2: peripheral oxygen saturation, FF: Fibro-Fatigue scale, HAMD: Hamilton Depression Rating scale, HAMA: Hamilton Anxiety Rating scale, Vaccination A,Pf,S: vaccination type either AstraZeneca (A), Pfizer-BioNTech (Pf) or Sinopharm (S), CRP: C-reactive protein

In this Table, we also examine whether the OSTOX/ANTOX ratio and CRP are predicted by SpO2 and body temperature while allowing for the effects of confounders. Regressions #7 and 8 display that the OSTOX/ANTIOX ratio was predicted by SpO2 and CRP by body temperature and male sex. **Figures 2** and **3** show the partial regressions of the OSTOX/ANTIOX ratio on SpO2 and of CRP on body temperature, respectively (after adjusting for age, sex, BMI, and TUD).

**Figure 2.**
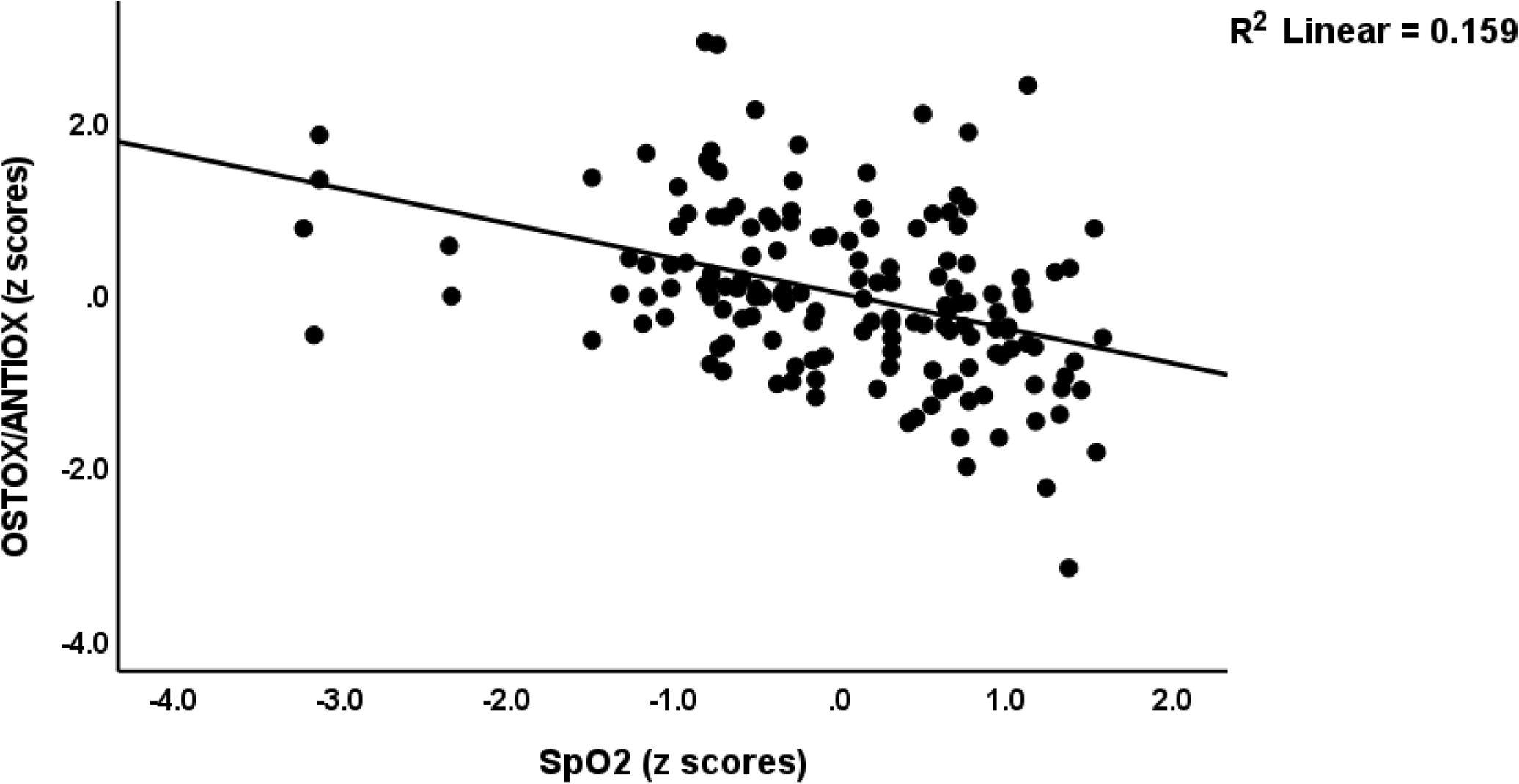
Partial regression of the oxidative stress toxicity / antioxidant (OSTOX/ANTIOX) ratio in Long COVID patients on peripheral blood oxygen saturation (SpO2) during the acute phase of COVID-19

**Figure 3.**
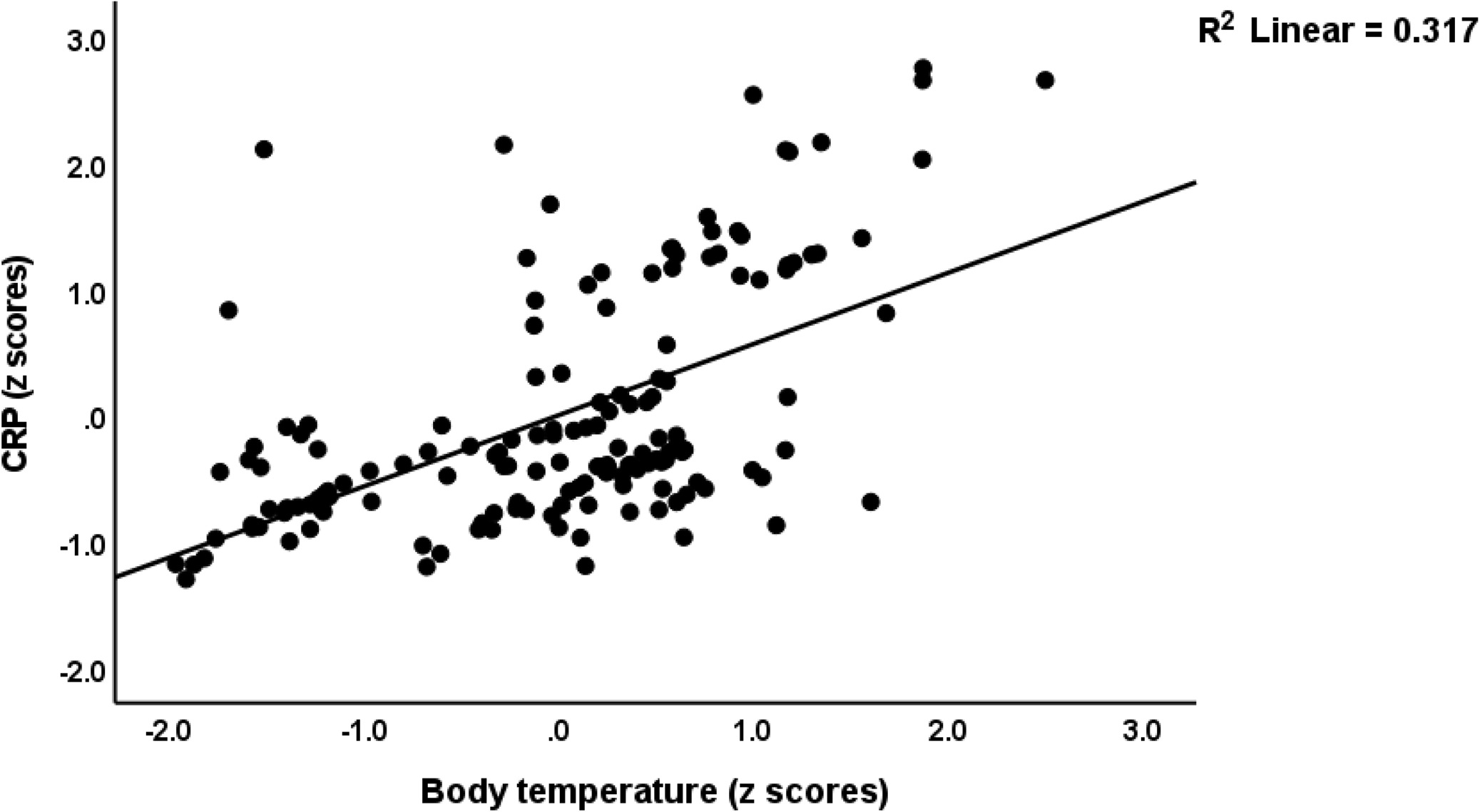
Partial regression of serum C-reactive protein (CRP) in Long COVID patients on peak body temperature (BT) during the acute phase of COVID-19

### Associations between physio-affective scores and all biomarkers combined

To examine the association between the combined effects of SpO2, body temperature and the biomarkers of Long COVID, we performed two types of analyses: a) Pearson’s correlation analyses between the physio-affective score and a new composite score computed as z body temperature - SpO2 + z OSTOX – z ANTIOX (dubbed the BTO2ONS index); and b) canonical correlation analyses with the physio-affective scores as dependent variables and SpO2, body temperature and a Long COVID biomarker composite score (z OSTOX + z CRP – z ANTIOX, dubbed the ONSCRP index) as explanatory variables.

Table 3 shows that the BTO2ONS index was significantly correlated with CRP and with all physio-affective scores in the total study group and the restricted study group of Long COVID patients (except for cognitive disorders). **Figure 4** shows the partial regression of the physio-affective score on the BTO2ONS composite score after controlling for age, sex, BMI, education and TUD.

**Figure 4.**
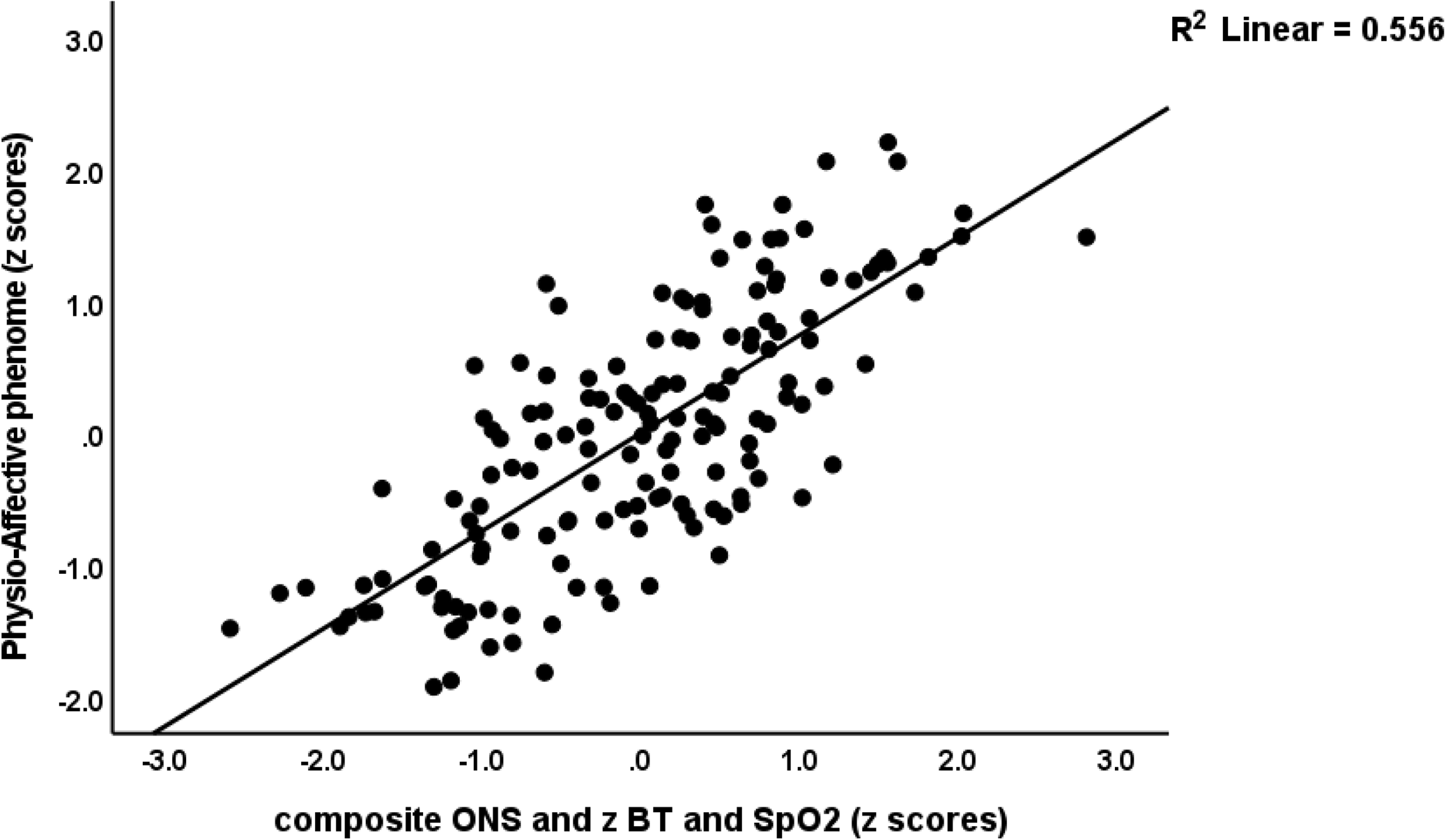
Partial regression of the physio-affective phenome score in Long COVID patients on a composite score based on increased oxidative stress toxicity and peak body temperature and reduced antioxidant defenses and peripheral oxygen saturation

**Table 6** shows the results of three canonical correlations with different sets of physio-affective scores as dependent variables. All CCAs showed that one canonical function was significant, while the other variates had no significant information about the variables. CCA #1 shows that the first canonical correlation between the biomarkers and total physio-affective scores was r=0.752. The first canonical variate based on SpO2, body temperature and the ONSCRP index explained 64.6% of the variance, indicating that this component is an overall measure of the adverse outcome pathways, and the first canonical variate based on the rating scale scores explained 89.4% of the variance indicating it is an overall measure of severity of the physio-affective phenome of Long COVID. Moreover, the amount of variance explained by the pathways in the total phenome was 50.6%. The same biomarker set explained 41.6% of the variance in a canonical variate extracted from pure FF and pure and physiosom HAMA and HAMD scores, and 39.1% in a canonical component based on autonomic and GIS symptoms, insomnia and chronic fatigue, while the canonical loading of cognitive disorder was lower (0.421).

**Table 6.**
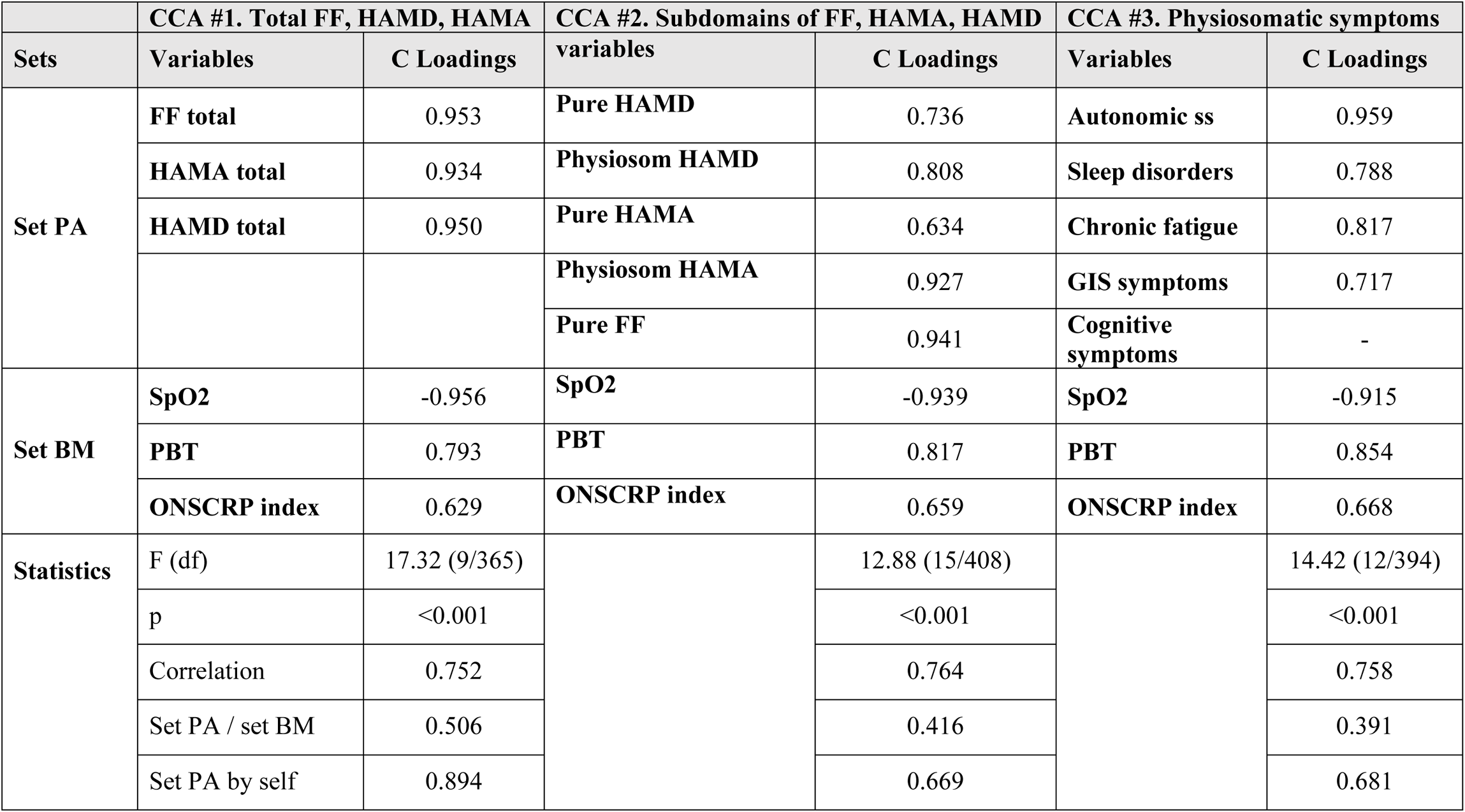
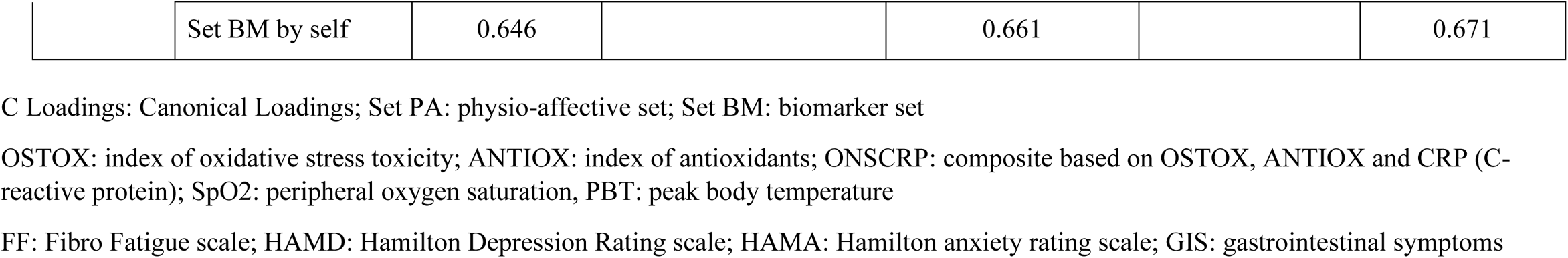
Results of canonical correlation analysis (CCA) with physio-somatic scores as dependent variables and biomarkers of acute and Long COVID-19 as explanatory variables

### Results of PLS and construction of a new endophenotypic class and a pathway phenotype

**Figure 5** shows a PLS model that examined whether the effects of SpO2 and body temperature during the acute phase of the physio-affective phenome of Long COVID (a latent vector extracted from the 5 rating scale subdomains) are mediated by IO&NS biomarkers. Therefore, we entered all ONS biomarkers and CRP as single indicators that could be predicted by SpO2 or body temperature, and whereby the ONS biomarkers, the overarching OSTOX/ANTIOX ratio and CRP were allowed to predict the physio-affective phenome, which was entered as a latent vector extracted from pure FF, and pure ad physiosom HAMA and HAMD scores. With an SRMR of 0.027, the model quality was adequate, and we found adequate values for construct reliability validity of the physio-affective phenome (AVE=0.731; composite reliability=0.931, Cronbach alpha=0.907, rho_A=0.920, all loadings > 0.777). Blindfolding revealed appropriate construct cross-validated redundancy for the physio-affective phenome LV (0.424). Non-significant paths and non-significant indicators are deleted from the model. The following are the findings of the complete PLS analysis (bias-corrected and accelerated using 5000 bootstraps,) as shown in Figure 5: a large part of the variance in the physio-affective phenome (60.3%) is explained by female sex, body temperature, SpO2 and OSTOX/ANTIOX ratio. The effects of SpO2 on the phenome score are in part mediated by effects on Gpx (t=-2.21, p=0.027) and NO (t=-2.05, p=0.041) which contribute to the OSTOX/ANTIOX ratio. Body temperature (t=4.02, p<0.001), SpO2 (t=-9.14, p<0.001), zinc (t=-2.74, p=0.006), TAC (t=-2.90, p=0.004), Gpx (t=-2.76, p=0.006), NO (t=2.75, p=0.006), MDA (t=2.85, p=0.004), and MPO (t=2.75, p=0.006), protein carbonyls (t=2.69, p=0.007) have significant total effects on the output latent vector.

**Figure 5.**
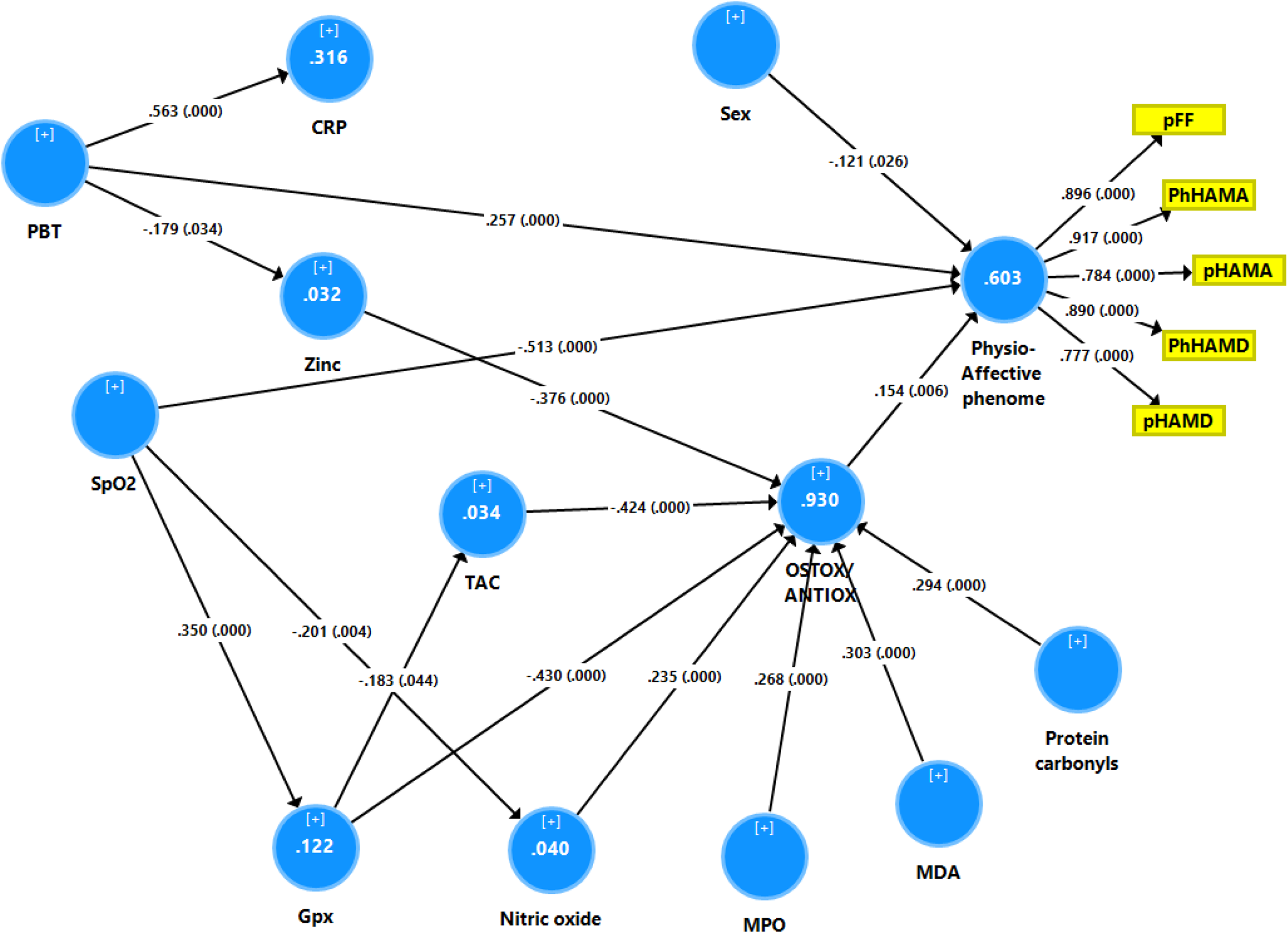
Results or Partial Least Squares (PLS) analysis which delineated that the effects of reduced oxygen saturation (SpO2) and increased peak body temperature (PBT) during the acute phase of COVID-19 on the physio-affective phenome of Long COVID are in part mediated by increased oxidative stress toxicity and lowered antioxidant defenses. The physio-affective phenome of Long COVID is entered as a latent vector extracted from subdomain scores on the Fibromylagia-Fatigue (FF), Hamilton Depression (HAMD) and Anxiety (HAMA) rating scales. pFF: pure physiosomatic FF symptoms; pHAMD and pHAMA: pure HAMD and HAMA scores, respectively; PhHAMD and PhHAMA: physiosomatic HAMD and HAMA symptom scores, respectively. CRP: C-reactive protein; Gpx: glutathione peroxidase; TAC: total antioxidant capacity; MPO: myeloperoxidase; MDA: malondialdehyde. Shown are path coefficients (with exact p values between brackets), loadings (with p-values) of the latent vector and explained variance (white figures in blue circles).

Based on all variables entered in this model (except sex) and the latent physio-affective score, we have performed a two-step cluster analysis with COVID infection as a categorical variable and all biomarkers as scale variables. **Figure 6** shows the features of the formed clusters. We discovered a three-cluster model with an appropriate measure of cohesiveness and separation of 0.52, consisting of a control cluster (n=36), patient cluster 1 (n=82) and patient cluster 2 (n-38). Figure 6 shows a clustered bar graph that was made using the significant indicators in the PLS analysis. Patient cluster 2 is a new endophenotype class of Long COVID characterized by very high body temperature, physio-affective scores, MDA, MPO, protein carbonyls and NO values, and very low SpO2, Gpx, zinc and TAC values as compared with cluster 1 (all p<0.01). We also constructed a new pathway phenotype by extracting a factor from ONSCRP, SpO2, PBT, and the physio-affective phenome (KMO=0.782, all loadings >0.753, explained variance=68.5%, Cronbach alpha=0.822).

**Figure 6.**
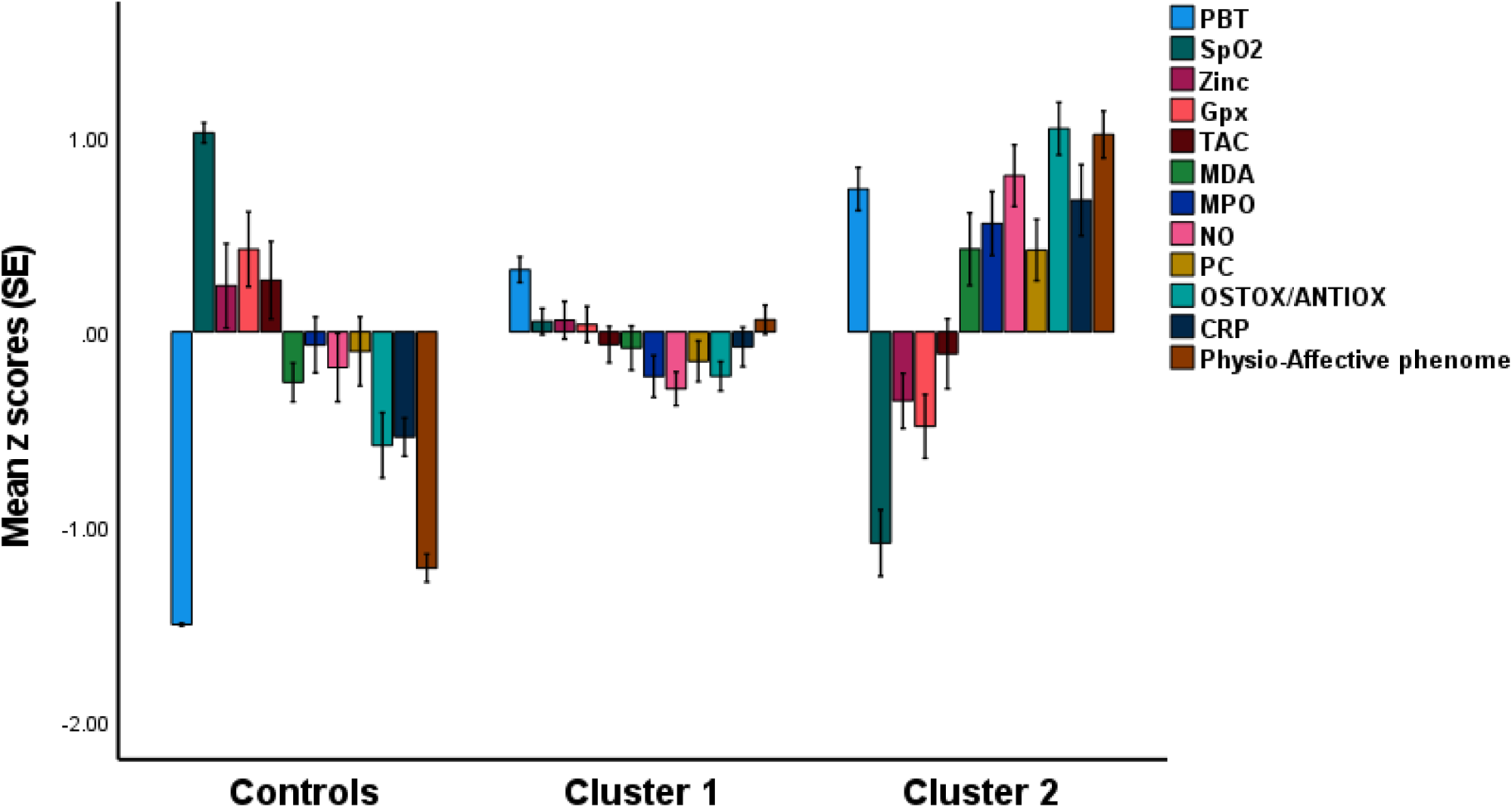
Results of cluster analysis with the formation of a new endophenotype class of patients with Long COVID (cluster 2) which is characterized by increased PBT (peak body temperature) and lowered peripheral oxygen saturation (SpO2) during acute COVID-19, and lowered zinc, Gpx (glutathione peroxidase) and TAC (total antioxidant capacity), and increased MDA (malondialdehyde), MPO (myeloperoxidase), NO (nitric oxide), PC (protein carbonyls), OSTOX/ANTIOX (oxidative stress toxicity / antioxidant defenses) ratio, CRP (C-reactive protein) and severity of the physio-affective phenome three to four months after the acute phase (Long COVID).

## Discussion

### Oxidative stress and immune activation in Long COVID

The current study’s first major finding is that a) Long COVID is associated with decreased antioxidant defenses, including Gpx, and a mild inflammatory process; and a) 31.7 percent of Long COVID patients belong to a cluster characterized by decreased SpO2, antioxidant levels, zinc, and Gpx, and increased body temperature, CRP, and OSTOX (increased MPO, NOx, MDA, and protein carbonyls). To put it another way, Long COVID is associated with immune-inflammatory processes and decreased antioxidant defenses, while a subgroup of Long COVID patients additionally exhibits increased NO production, oxidative damage to proteins (protein carbonyls) and lipids with increased aldehyde formation (MDA), and immune-oxidative stress response (increased MPO and CRP).

Previous research found that between 1.8 and 24.5 percent of Long COVID patients had elevated CRP levels ^11, 49–52^, which were operationalized as >10 mg/L or >2.9 mg/dL. Other authors found that median CRP levels varied between 0.6 and 2.9 mg/L ^4, 53–55^. Nevertheless, in the present study, we detected that the patient group with Long COVID showed increased CRP levels compared with control levels, indicating a mild inflammatory response. Previously, a meta-analysis reported that 13 out of 14 studies showed increases in at least one measure of inflammation either in a subgroup of patients or throughout the whole post-COVID period ^56^. In addition, more CD4+ T cells expressed interferon-γ and interleukin (IL)-2, and increased plasma levels of IL-6, macrophage inflammatory protein 1, IL-1β, tumor necrosis factor-a and the soluble IL-1 receptor antagonist in the Long COVID patients ^49, 52, 53, 57–60^, indicating activation of M1 macrophages and T helper (Th)-1 phenotypes ^33^. Mehandru and Merad reviewed that Long COVID is accompanied by elevated neutrophils, lipid abnormalities, and reduced serum albumin ^61^.

There are some data that acute COVID-19 infection is accompanied by signs of oxidative stress, including increased expression of ROS-response genes and glycolysis in peripheral blood mononuclear cells ^62^, increased MDA (TBARS assay), F-isoprostane, 8-hydroxy-2’-deoxyguanosine, AOPP and nitrotyrosine levels, and lowered glutathione (GSH), −SH groups, selenium, zinc, TAC and vitamin D ^39, 63–69^. Lage et al. reported that activation of the NLRP3 inflammasome, including increased mitochondrial superoxide production and lipid peroxidation, may persist after short-term patient recovery ^70^.

As such, our work demonstrates that the oxidative stress biomarkers during Long COVID are quite similar to those found during the acute phase, except for AOPP, nitrotyrosine and TAC levels, which were altered in acute COVID-19 but not in Long COVID. Nonetheless, our study demonstrates that a persistent imbalance between oxidative stress toxicity and reduced antioxidant defenses is a key component of Long COVID and that increased aldehyde, neutrophil-associated MPO, protein carbonyl, and NO production are the major neurotoxic pathways, which, when combined with decreased antioxidant defenses, may have a variety of detrimental effects as reviewed before ^71^.

Given that the Long COVID group as a whole showed Gpx deficiency, while only a subset of patients demonstrated oxidative damage, we may infer that Gpx deficiency is the main cause of the consequent lipid and protein damage. Gpx has been shown to degrade lipid hydroperoxides to their respective alcohols and free hydrogen peroxide to water ^72, 73^, hence reducing oxidative damage ^71^. Due to their negative impact on the vascular endothelium and inflammatory processes, these activated oxidative stress pathways contribute to the pathophysiology of COVID-19 and Long COVID ^74–76^. Additionally, MPO generated by neutrophils during inflammation may result in irreversible protein and lipid alterations and increased levels of oxidized low-density lipoprotein, thereby promoting atherogenesis ^77^. Furthermore, the resulting chlorinative stress may lead to multiorgan damage in COVID-19 through various mechanisms, including oxygen competition for heme-binding sites, decreased oxygen saturation, hemoglobin-heme iron oxidation, heme degradation, and iron release ^78^. Although NO production was enhanced considerably in Long COVID, nitrotyrosine levels were not altered, although increased nitrotyrosine indicates an increase in reactive nitrogen species with increased NO2 binding to tyrosine, thereby forming an immunogenic neoepitope ^79^. Nonetheless, future research should explore if the increased NO generation in Long COVID results in nitrosative stress associated with hypernitrosylation (increased NO or nitroso binding), which has a variety of neurotoxic consequences ^80^. It is critical to remember that low zinc levels have a wide impact on the immune system ^81, 82^ and that zinc itself may reduce viral replication, including SARS-CoV-2 replication ^83^.

### Biomarkers of acute COVID-19 predict IO&NS biomarkers of Long COVID

The second major finding of this study is that decreased SpO2 levels during the acute phase of COVID-19 significantly predict oxidative toxicity, increased NO production, and decreased Gpx and antioxidant defenses, whereas increased peak body temperature levels during the acute phase predict increased CRP and decreased zinc and antioxidant levels in Long COVID. As a result, both decreased SpO2 and higher body temperature contribute to Long COVID’s increased OSTOX/ANTIOX ratio.

As mentioned in the Introduction, both lowered SpO2 and increased body temperature are part of the immune-inflammatory response, predict critical illness and death, and are manifestations of the infectious-immune-inflammatory core of acute COVID-19 ^84^. SpO2 is often lowered in COVID-19, particularly in more severe instances and in the presence of CCTAs ^85, 86^, and both CCTAs and decreased SpO2 are highly related to immunological activation ^25^. COVID-19-associated microvascular damage, such as endothelitis, microthrombosis, capillary damage, damage to pericytes, and fluid buildup in the alveoli, results in hypoxia and reduced SpO2 ^87^. Prolonged systemic hypoxia has been shown to affect the redox equilibrium and induce oxidative stress ^88, 89^. These effects drive inflammatory processes and nitro-oxidative pathways ^90^ and the resultant excess of reactive oxygen species may induce oxidative damage, aggravating systemic tissue damage and contributing to more severe COVID-19 ^91^. COVID-19 individuals who require invasive mechanical breathing, admission to an intensive care unit, or lengthy hospitalization are more likely to sustain long-term tissue damage as a result of persistent symptoms ^92–94^. As such, it appears that COVID-19 (and other viral infectious diseases) is accompanied by three different inflammatory phases: a) beneficial protective inflammation, which combats the infection and may mount a restorative repair, b) hyperinflammation, which may lead to a cytokine storm and critical disease or even death, and c) non-resolving inflammation resulting in a protracted mild inflammatory state ^95^ as observed in Long COVID. All in all, our results show that the severity of the infectious-immune-inflammatory response during acute COVID-19 may, at least in part, predict mild inflammation and increased nitro-oxidative damage in Long COVID.

### IO&NS biomarkers predict the physio-affective phenome of Long COVID

The third and most noteworthy finding of this study is the discovery of a) a pathway phenotype comprising SpO2, body temperature, OSTOX, ANTIOX, CRP and the physio-affective phenome; and b) an endophenotype class of Long COVID patients (31.7%) who show simultaneously severe abnormalities in SpO2 and peak body temperature during acute COVID-19, and an increased OSTOX/ANTIOX ratio, CRP and increased total HAMD, HAMA and FF total and subdomain scores during Long COVID. The changes in the rating scale scores indicate the presence of moderate depression and anxiety symptoms consistent with major depression and generalized anxiety disorder and the presence of a chronic fatigue syndrome (lasting 2-3 months), including chronic fatigue, insomnia, autonomic and gastro-intestinal symptoms, and, to a lesser extent, cognitive impairments. Additionally, approximately 60% of the variance in the severity of the physio-affective phenome is explained by the cumulative effects of decreased SpO2 and increased peak body temperature (thus, the severity of the acute infectious phase) coupled with increased CRP and OSTOX/ANTIOX ratio. Most notably was the impact of MDA and NO and decreased Gpx and zinc, although protein carbonyls and MPO, as well as decreased TAC, all contribute to the physio-affective phenome.

According to a recent meta-analysis, the percentage of people suffering chronic fatigue twelve weeks or more after exposure to COVID-19 was 0.32, whereas cognitive impairment was 0.22 ^56^. Additionally, a narrative synthesis indicated that a subgroup of Long COVID patients had elevated proinflammatory indicators and significant functional impairment. Another meta-analysis found that 80 percent of patients infected with SARS-CoV-2 had one or more long-term symptoms and that the top most typical symptoms were chronic fatigue (58%), headache (44%), attention problems (27%), hair loss (25%), and dyspnea (24%), although memory loss, anxiety, depression, insomnia, digestive disorders, and autonomic symptoms were also widespread (from 10-20 percent) 15–110 days after virus infection ^96^. Mehandru and Merad reviewed the many clinical manifestations of long-haul COVID, including systemic and musculoskeletal manifestations (chronic fatigue and fibromyalgia symptoms), neuropsychiatric symptoms (insomnia, depression, anxiety, neurocognitive deficits), autonomic and cardio-vascular, GIS and renal and respiratory manifestations ^61^. According to some other studies, 50 to 98 percent of the Long COVID patients complain of fatigue as the primary symptom ^1, 12, 97^. Additionally, mood abnormalities are common ^2^, particularly despair and anxiety ^23^, whereas 58.4 percent of Long COVID patients have cognitive impairment and 34–51-78% have memory problems, particularly memory loss ^2, 18, 22, 94, 98^. Additionally, other studies indicate that dyspnea, autonomic symptoms, post-traumatic stress and fibromyalgia symptoms (muscle aches, myalgia, and articular pain) are common ^99–101^. Notably, the severity of acute COVID-19 infection affects the development of neuropsychiatric symptoms, with ICU survivors being more likely (56%) than non-ICU survivors to develop a neuropsychiatric condition ^24^. Most importantly, the results of our study show that chronic fatigue, depression and anxiety symptoms in Long COVID are all manifestations of the same physio-affective core, which is strongly associated with OSTOX/ANTIOX and inflammatory signs.

There is now evidence that major depression, generalized anxiety disorder and chronic fatigue syndrome are IO&NS-related disorders characterized by increased levels of inflammatory mediators, including CRP, oxidative damage to lipids with increased aldehyde formation, damage to proteins, increased NO production and hypernityrosylation and lowered antioxidant levels including zinc, GSH and Gpx and total antioxidant capacity ^28, 31, 71, 102^. We have reviewed previously the mechanistic explanations of how these multiple IO&NS pathways cause neuroaffective toxicity and, consequently, the onset of chronic fatigue syndrome and physiosomatic and affective symptoms ^28, 31, 71, 102–104^. All in all, the hyperinflammation that may accompany SARS-CoV-2 infection is not only associated with severe lung pathology ^105, 106^, a poorer clinical outcome ^105, 107^ and higher mortality ^108^ but also with increased OSTOX/ANTIOX and physio-affective symptoms three to four months later. It should be stressed that only part of the variance in Long COVID physio-affective symptoms was explained by IO&NS pathways indicating that other mechanistic processes are involved. In this respect, it is plausible that autoimmune responses, which play a role in chronic fatigue syndrome and depression ^28, 109^ and Long COVID ^110^, and pulmonary fibrosis with scarring and cardiac remodeling ^61^ contribute to the physio-affective phenome.

Finally, it is also important to note that vaccinations with AstraZeneca (viral vector, genetically modified virus vaccine) and Pfizer (mRNA vaccine), but not Sinopharm (inactivated virus vaccine), may aggravate the physio-affective phenome and, in particular, the physiosomatic symptoms of the HAMA and HAMD subdomains. It is known that this type of corona vaccine may cause Long Covid–like symptoms, including anxiety, depression and fatigue, T cell activation, autoimmune responses, increased production of spike protein, and impairments in type 1 interferon signaling111, 112.

## Limitations

This research would have been more interesting if we had also assayed the cytokine network, the NLRP3 inflammasome and biomarkers of hypernitrosylation. Follow-up studies should examine the same rating scales and biomarkers at later time points, including 6 months, one and two years after the acute infection, to delineate which patients show protracted chronic fatigue syndrome and GAD (which tend to be chronic disorders) and remitted depression (which most often comes in short-lasting episodes) ^44, 95^.

## Conclusions

Around 60 percent of the variation in the physio-affective phenome of Long COVID is explained by OSTOX/ANTIOX, peak body temperature and SpO2. Lowered SpO2 predicts the alterations in Gpx and NO production during Long COVID, while increased body temperature predicts increased CRP and reduced ANTIOX and zinc levels in Long COVID. The impact of the immune-inflammatory response during acute COVID-19 on the physio-affective phenome of Long COVID is partially mediated by OSTOX/ANTIOX. Post-viral physio-affective symptoms have an inflammatory origin and are partially mediated by neuro-oxidative damage and lowered antioxidant defenses.

## Author**’**s contributions

HTA and DSA recruited the patients and collected blood samples. HTA and HAH measured the serum biomarkers. MM performed the statistical analysis. All authors contributed to the drafting and editing of the manuscript, and all authors approved the final version of the manuscript.

## ETHICS APPROVAL AND CONSENT TO PARTICIPATE

The research was approved by the University of Kufa’s institutional ethics board (8241/2021) and the Najaf Health Directorate-Training and Human Development Center (Document No.18378/ 2021).

## HUMAN AND ANIMAL RIGHTS

The study was conducted according to Iraq and international ethics and privacy laws and was conducted ethically following the World Medical Association Declaration of Helsinki. Furthermore, our IRB follows the International Guideline for Human Research protection as required by the Declaration of Helsinki, The Belmont Report, CIOMS Guideline and the International Conference on Harmonization in Good Clinical Practice (ICH-GCP).

## CONSENT FOR PUBLICATION

All participants gave written informed consent before participating in this study.

## AVAILABILITY OF DATA AND MATERIALS

The dataset generated during and/or analyzed during the current study will be available from the corresponding author (M.M.) upon reasonable request and once the dataset has been fully exploited by the authors.

## Acknowledgments

We thank the staff of Al-Sadr Teaching Hospital and Al-Amal Specialized Hospital for Communicable Diseases in Najaf governorate-Iraq for their help in collecting samples. We also thank the high-skilled staff of the hospitals’ internal labs for their help in estimating biomarkers levels.

